# Surgical Decision-Making in Neonates with Borderline Left Ventricular Hypoplasia without Significant Aortic or Mitral Valve Stenosis using Cardiovascular Magnetic Resonance

**DOI:** 10.1101/2024.05.31.24308306

**Authors:** Aswathy Vaikom House, Aakansha Singh, Ankavipar Sapruangrang, Edgar Jaeggi, David J. Barron, Christopher Z. Lam, Shi-Joon Yoo

## Abstract

**Background:** Most patients with significant left ventricular (LV) hypoplasia undergo single ventricle (SV) palliation, but biventricular (Bi-V) repair is viable in some patients with borderline LV hypoplasia. We sought to identify CMR (cardiovascular magnetic resonance) criteria predictive of successful primary Bi-V repair in neonates with borderline LV hypoplasia without significant stenosis of the mitral valve (MV) and aortic valve (AV), and to determine reasons for reintervention after successful Bi-V repair.

**Methods:** This retrospective study included all patients with borderline LV hypoplasia who underwent CMR from 2003-2024 for surgical decision-making. Patients with abnormal segmental connections, atrioventricular septal defects, unrestrictive ventricular septal defects, those with more than mild MV stenosis (mean Doppler flow gradient > 5 mmHg) and/or more than mild AV stenosis (peak Doppler flow gradient >20 mmHg) were excluded. Patients were divided into two groups based on initial intervention - primary Bi-V repair and hybrid/ other staging procedure. Outcomes were categorized as successful primary Bi-V repair, successful staged Bi-V repair and failure to achieve Bi-V repair (hybrid followed by SV palliation, transplant, death). Fisher exact test and Mann-Whitney U test was utilized to explore potential relationships. ROC curves were used to test diagnostic accuracy of parameters to predict successful primary Bi-V repair.

**Results:** Among 37 patients meeting the inclusion criteria, 23 (62%) patients underwent successful primary Bi-V repair, 8 (22%) underwent staged Bi-V repair, 6 (16%) failed to achieve Bi-V repair. Patients who underwent successful primary/ staged Bi-V repair had higher values for left ventricular diastolic volume index (LVEDVi 28 mL/m^2^ vs. 17.4.00 mL/m^2^; p <0.002), higher blood flow volume through the ascending aorta (Q_Ao_:1.99 L/min/m^2^ vs. 0.97 L/min/m^2^, p <0.012), and Q_Ao_ / superior vena cava (Q_SVC_) flow ratio (1.44 vs. 0.85, p =0.034) compared to those who had failure to achieve Bi-V repair. CMR LVEDVi cutoff of CMR 27 mL/m², had 87% sensitivity and 79% specificity with an AUC of 87.6% and Q_Ao_ threshold of 1.9 L/min/m^2^ had 65.2% sensitivity and 92.9% specificity (AUC: 86.0%) to predict successful primary Bi-V repair. Of 31 patients with primary or staged Bi-V repair, 7 (22%) underwent reinterventions for LVOT obstruction followed by mitral stenosis.

**Conclusions:** CMR plays a critical role in pre-operative evaluation, surveillance and decision-making in patients with borderline LV hypoplasia. In patients with borderline LV hypoplasia without MV/AV stenosis, successful primary Bi-V repair can be achieved when the CMR-derived LVEDVi is >27 mL/m^2^ and Q_Ao_ is > 1.99 L/min/m^2^.

## 1. Introduction

Borderline left ventricle is a heterogenous condition characterized by a spectrum of underdevelopment of the left ventricle with variable degrees of stenosis and/or hypoplasia of the aortic valve (AV) and/or mitral valve (MV) as well as hypoplasia of the ascending aorta and aortic arch.^1-5^ In these patients, the decision for bi-ventricular (Bi-V) and single ventricle (SV) repair is crucially important .^3–5^ While Bi-V repair is attractive,^6^ it does carry significant risks: failed Bi-V strategy is associated with high mortality and morbidity, and SV conversion after a failed Bi-V repair is a high-risk operation with suboptimal outcomes.^7,8^ Therefore, robust decision-making criteria for Bi-V versus SV repair are crucial.

It is important to recognize that the same criteria cannot be applied to all patients with borderline LV hypoplasia. There are two distinct groups of patients with LV hypoplasia: those with increased LV afterload as seen in patients with critical aortic stenosis^9,10^ often with concomitant MV stenosis, and those with decreased LV preload, typically due to aortic arch obstruction and a widely patent ductus arteriosus in the newborn period. A distinction must be drawn to the latter group, where there is borderline LV hypoplasia, with *hypoplasia* but without significant stenosis of the MV and AV. This group of patients is notable because their LV has not faced increased in-utero pre- and afterload. Unlike patients with critical AV stenosis, where the LV is elliptical, tense, often with endocardial fibroelastosis where the interventricular septum bows toward the RV and has limited growth potential, the latter group of patients have a crescentic LV with the septum bowing toward the LV, healthy myocardium and the potential to grow once preload increases and right ventricular pressure declines (Figure 1). Tchervenkov et al introduced the term “hypoplastic left heart complex (HLHC)”^11–13^ to describe this group of patients with LV hypoplasia and hypoplasia of the MV and AV without significant stenosis, and described high rate of successful Bi-V repair in this group. More recently, others have proposed criteria for Bi-V repair in this specific group of patients with borderline LV hypoplasia primarily based on echocardiographic measurements.^14,15^ Over the last two decades, cardiac magnetic resonance (CMR) has emerged as the gold-standard technique to measure ventricular volumes.

**Figure 1.**
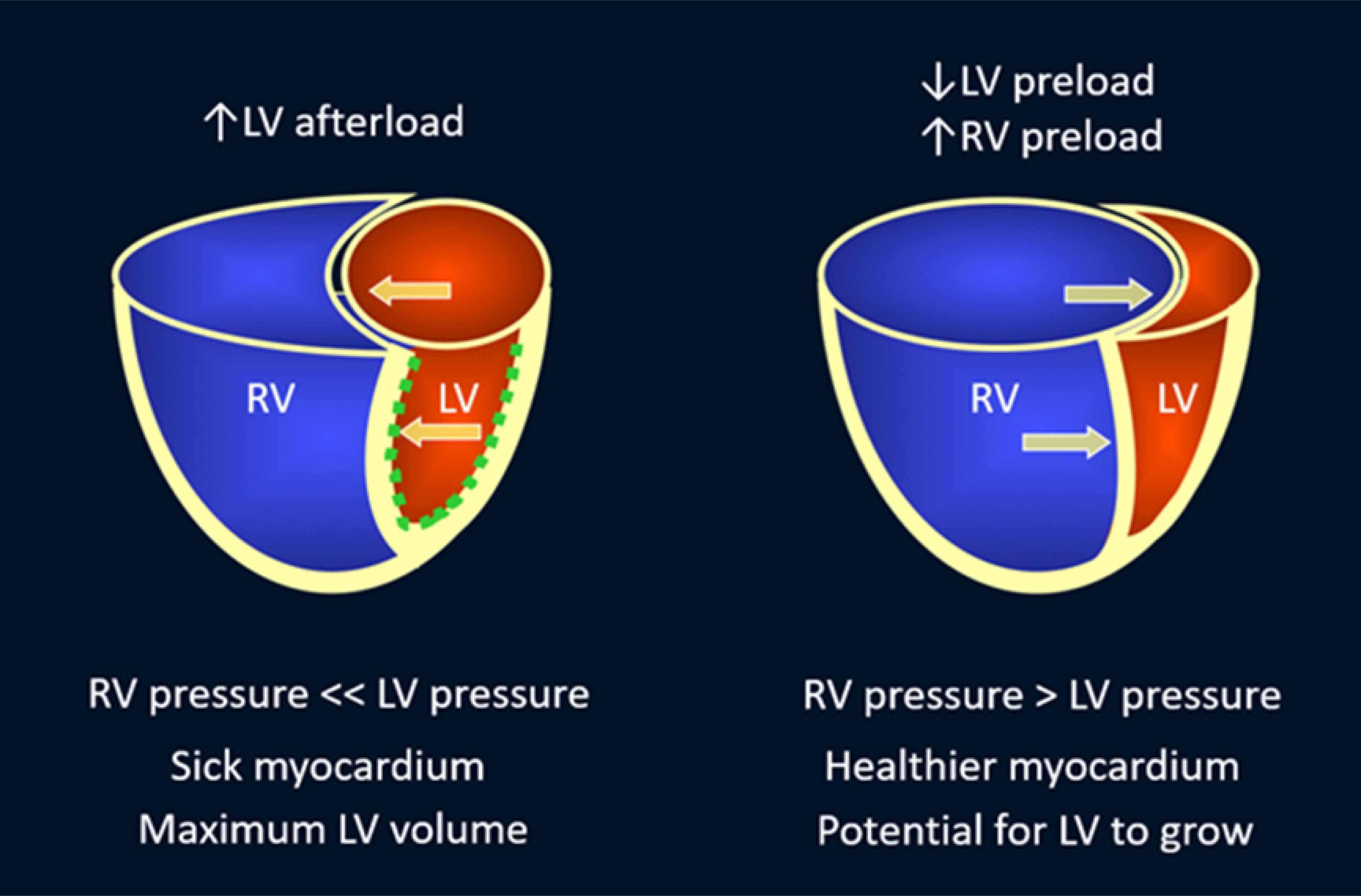
Schematic showing differences between left ventricular (LV) hypoplasia with increased LV afterload as in critical aortic stenosis and LV hypoplasia with decreased LV preload as in so-called hypoplastic left heart complex. In the former, the ventricular septum is bowing toward the right ventricle (RV). The LV is hypertensive and hypertrophic, and globular or elliptical. The myocardium is damaged and frequently lined with endocardial fibroelastosis (dotted green line). In the latter, the ventricular septum is bowing toward the LV. The LV is crescentic in shape and the myocardium is neither hypertrophic nor damaged.

In this retrospective study, we aimed to identify CMR-derived criteria predictive of successful primary Bi-V repair in neonates with borderline LV hypoplasia without significant MV or AV stenosis. We also sought to determine the reasons for reintervention after an initially successful Bi-V repair.

## 2. Methods

This retrospective study was approved by the Institutional Research Ethics Board. Using the institutional CMR database, we identified all patients with an initial echocardiographic diagnosis of borderline LV hypoplasia who underwent CMR for assessment of the ventricular volumes and hemodynamics for a surgical decision between January 2003 and April 2024.

Excluded from the study were patients with discordant atrioventricular and/or ventriculoarterial connections, atrioventricular septal defects, unrestrictive ventricular septal defects, or anomalous pulmonary venous connections. We further reviewed the initial echocardiogram and excluded patients who displayed more than mild MV stenosis (mean inflow Doppler flow gradient > 5 mmHg) and/or more than mild AV stenosis (peak Doppler flow gradient >20 mmHg). Patients with structurally abnormal valves such as bicuspid AV and parachute MV were included if the baseline MV/AV stenosis were only mild.

Patients were divided into two groups based on the initial surgical and/or catheter-based interventions as followed:

- Patients who underwent primary complete Bi-V repair
- Patients who underwent hybrid or other staging procedure

Outcomes were categorized as

- <u>Successful primary Bi-V repair</u> – initial complete repair with transplant-free survival at most recent follow up;
- <u>Successful staged Bi-V repair</u> – initial hybrid / staging procedure followed by complete Bi-V repair with transplant-free survival at most recent follow up;
- <u>Failure to achieve Bi-V repair</u> – initial hybrid/ staging procedure followed by conversion to SV palliation, death or transplantation.

### Data Collection

Clinical, diagnostic (echocardiograms, CMR, cardiac catherization), and catheter-based and surgical procedural data were reviewed. Clinical data collected included patient demographics, characteristics and outcome, including the need for transplantation and death. The initial neonatal echocardiogram, follow-up echocardiogram prior to definitive surgery (Primary Bi-V repair, Staged Bi-V repair or SV conversion) and last available echocardiograms were reviewed.

Echocardiographic measurements were performed from offline recordings by a single experienced observer (EJ). MV and AV annulus measurements were made in the 4-chamber and parasternal long-axis views, respectively. Left ventricular end-diastolic volume index (LVEDVi) was measured in the apical 4-chamber views. Left ventricular end-diastolic dimension (LVEDD) was measured in the parasternal short axis view. The ventricular volumes and valve diameters were indexed to the patient’s body surface area (BSA) using the z-scores of the published nomograms ^16^. The valvular function and morphology were recorded.

CMR was performed on a 1.5 T scanner (Avanto Fit, Siemens Healthineers, Erlangen, Germany) using a feed-and-sleep technique^17^ or under general anesthesia. The CMR protocol consisted of cine imaging in axial, 2-chamber, 4- chamber, and short-axis planes, through-plane phase-contrast imaging of the main, right, and left pulmonary arteries, ascending aorta at the level of the right pulmonary artery, descending aorta at the diaphragm, patent ductus arteriosus, superior vena cava, and atrioventricular valves, and contrast-enhanced angiography. Late gadolinium enhancement (LGE) was performed in 31 patients. Ventricular volumes and flow volumes were measured using a commercial software package (Cardiac Function Analysis and Flow Analysis; Medis Medical Imaging, Leiden, The Netherlands).

### Statistical Methods

Continuous data was presented as median with interquartile range (IQR), while for categorical data we used numbers and percentages. The normality of continuous variables was assessed using the Shapiro-Wilk test before analysis. For categorical variables, the Fisher exact test was utilized to explore potential relationships, while the Mann-Whitney U test was applied to assess differences across groups in continuous variables. Additionally, diagnostic test accuracy for different cardiac indices and their association with successful primary Bi-V repair was evaluated, where optimal cut-off points were determined for selected variables. Sensitivity, specificity, and the area under the curve (AUC) were calculated to ascertain the diagnostic performance of these markers. The Kaplan–Meier curve and Cox proportional hazards model were used to analyze time to death/transplantation and freedom from reintervention. Data analysis was done with Rstudio (Version 4.2.2). Throughout, a significance threshold of p < 0.05 was maintained.

## 3. RESULTS

### 3.1 Baseline Characteristics

Our study included a total of 37 patients. The median birth weight and gestational age presentation was 3.29 (3.00-3.55) kg and 39 (38.0-39.6) weeks, respectively for the entire cohort. Out of 37 patients with borderline LV hypoplasia meeting the criteria, 23 underwent primary biventricular repair and all showed transplant-free survival at the last follow-up. There were 15 patients with morphologically abnormal MV – (parachute MV: n=12: dysplastic MV: n=2; double orifice MV: n=1). 19 patients had a bicuspid aortic valve. None of the 31 patients who underwent LGE showed abnormal myocardial enhancement. There were no significant differences in baseline findings between patients who underwent successful primary Bi-V repair vs those who underwent initial hybrid/staging operation ***(Table 1)***. Of the 14 patients with an initial hybrid or staging procedure, 8 proceeded to a staged Bi-V repair, while 6 failed to achieve a Bi-V circulation ***(Figure 2)***.

**Figure 2.**
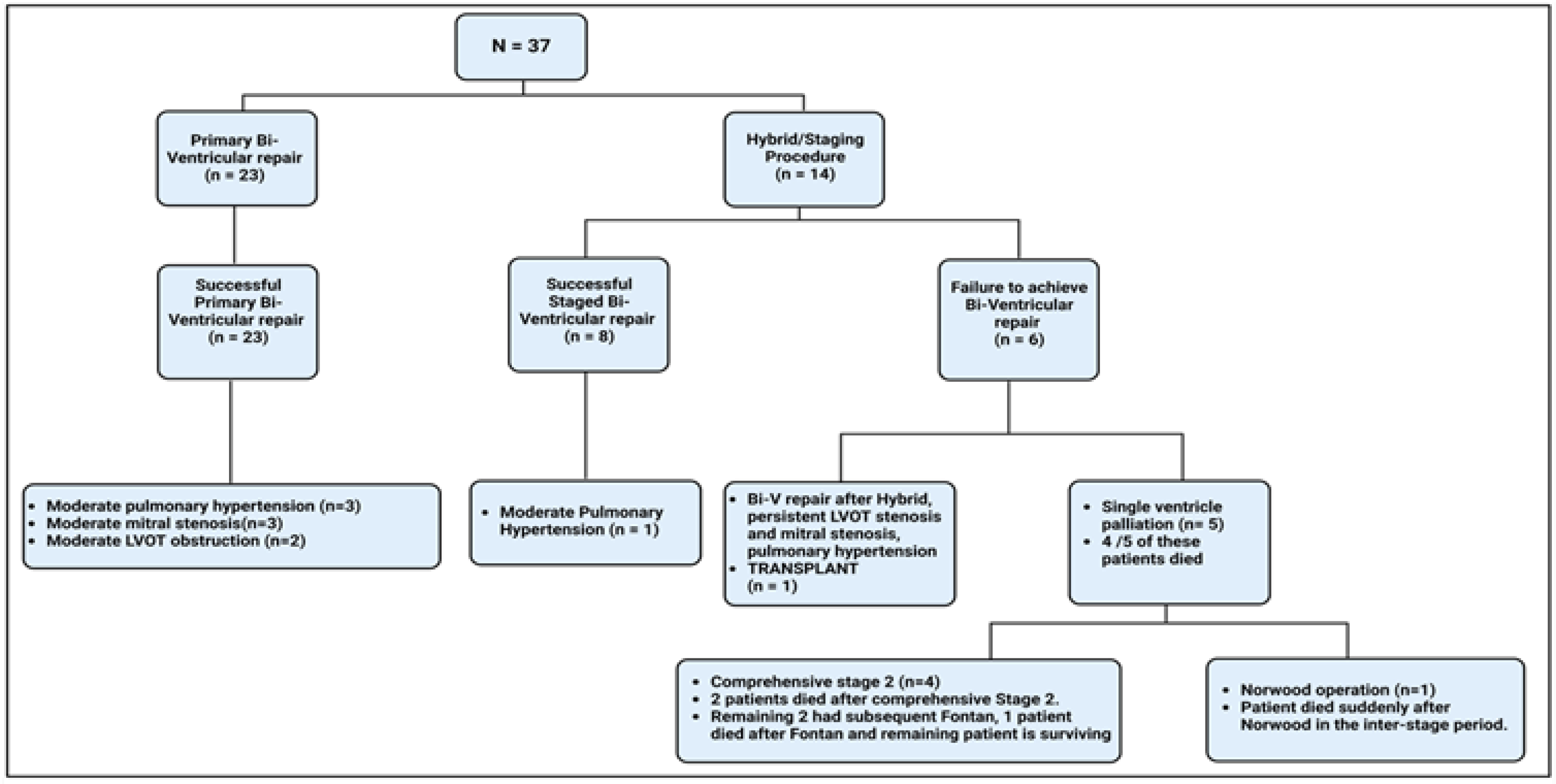
Summary of initial intervention and outcomes of study cohort.

**Table 1.** Baseline summary of included patients.

| Variable | Level | Overall | Bi-V Repair as Initial Operation<br>n = 23 | Hybrid Initial Operation<br>n = 14 | P-value |
| --- | --- | --- | --- | --- | --- |
| Gender (%) | Female | 15 (40.5) | 10 (43.5) | 5 (35.7) | 0.903† |
|  | Male | 22 (59.5) | 13 (56.5) | 9 (64.3) |  |
| Morphologically abnormal valves (%) | Absent | 14 (37.8) | 8 (34.8) | 6 (42.9) | 0.887† |
|  | Present | 23 (62.2) | 15 (65.2) | 8 (57.1) |  |
| Bilateral superior venacava (%) | Absent | 25 (67.6) | 15 (65.2) | 10 (71.4) | 0.977† |
|  | Present | 12 (32.4) | 8 (34.8) | 4 (28.6) |  |
| Mortality transplant (%) | No | 32 (86.5) | 22 (95.7) | 10 (71.4) | 0.111† |
|  | Yes | 5 (13.5) | 1 (4.3) | 4 (28.6) |  |
| Birth weight kg (median [IQR]) |  | 3.29 [3.00, 3.55] | 3.33 [3.05, 3.53] | 3.23 [2.90, 3.57] | 0.685§ |
| Gestational age in weeks (median [IQR]) |  | 39.00 [38.00, 39.60] | 39.00 [38.00, 39.60] | 38.50 [38.00, 39.53] | 0.824§ |
| Age at examination in days (median [IQR]) |  | 2.00 [1.00, 5.00] | 2.00 [1.00, 5.00] | 3.00 [2.00, 4.00] | 0.68§ |
| Height in cm (median [IQR]) |  | 50.00 [48.00, 51.00] | 49.00 [46.00, 50.00] | 50.00 [49.00, 51.75] | 0.14§ |
| Weight in kg (median [IQR]) |  | 3.30 [3.00, 3.68] | 3.30 [3.00, 3.64] | 3.30 [2.89, 3.68] | 0.9§ |
| Body surface area in m<sup>2</sup> (median [IQR]) |  | 0.21 [0.20, 0.22] | 0.20 [0.20, 0.21] | 0.21 [0.20, 0.22] | 0.091§ |

### 3.2 Imaging features associated with successful primary Bi-V repair

#### 3.2.1 CMR Parameters

Patients who underwent successful primary Bi-V repair exhibited higher values for LVEDVi (29 mL/ m^2^ vs. 20.00 mL/m2; p <0.001), LVSVi (17 ml/m2 vs. 11.70 ml/ m^2^; p=0.001), higher blood flow volume through the ascending aorta (Q_AO_: 2.1 L/min/ m^2^ vs. 1.27 L/min/ m^2^, p <0.001), and ascending aorta / superior vena cava (Q_SVC_) flow ratio (1.49 vs. 0.98, p =0.027) compared to those who initially underwent hybrid/staging procedure (***Table 2, Figure 3***).

**Figure 3.**
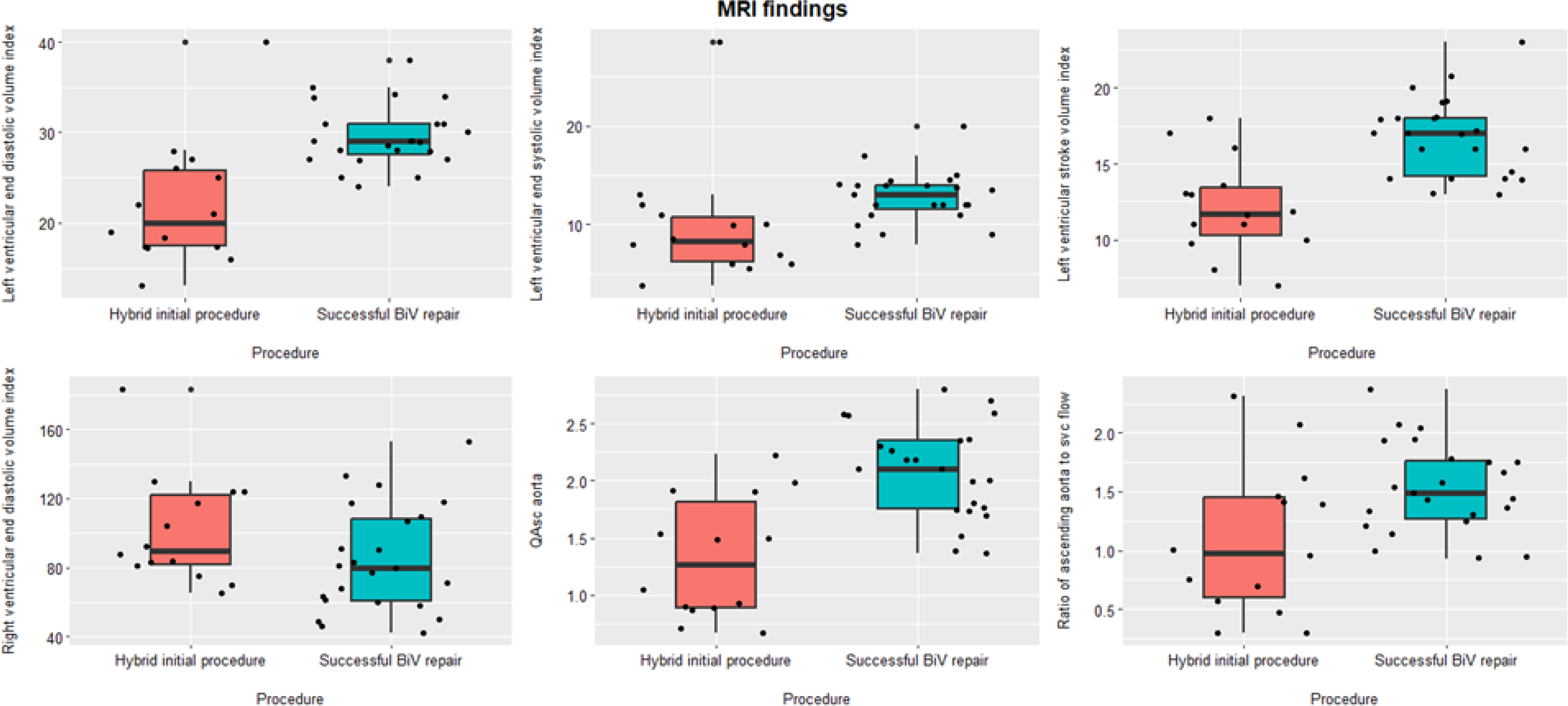
Comparison of MRI characteristics between primary Bi-V repair and hybrid group.

**Table 2.**
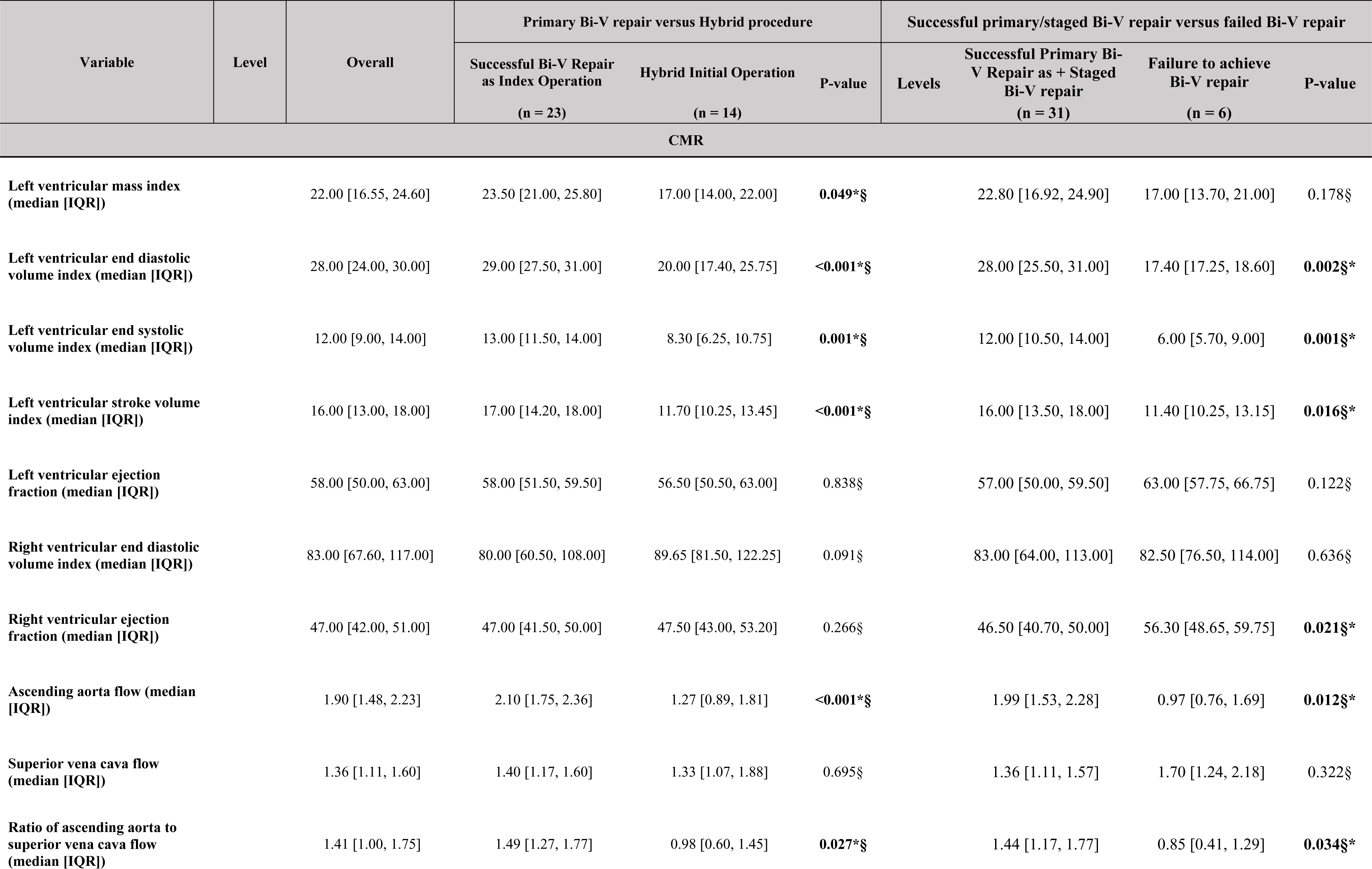
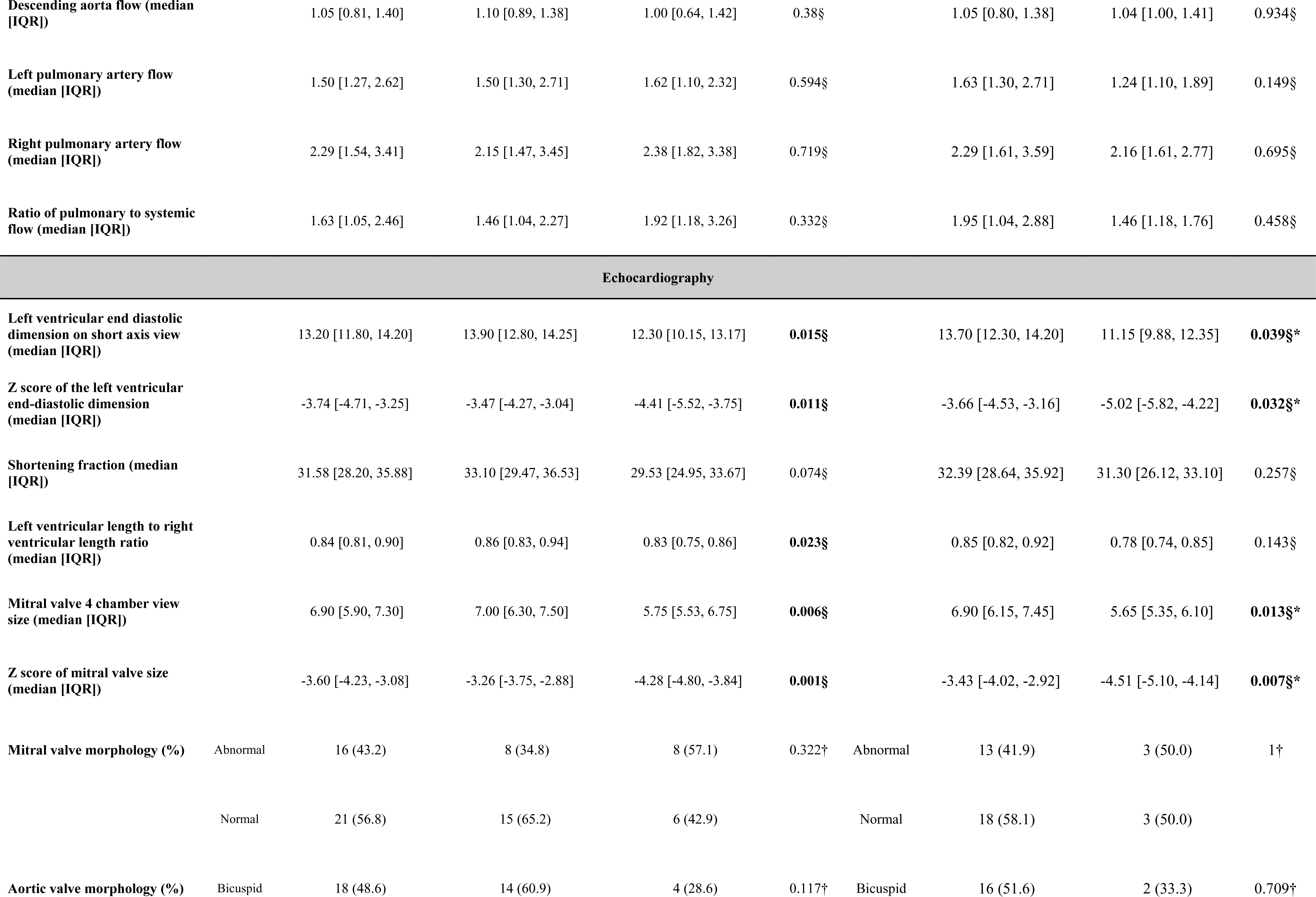
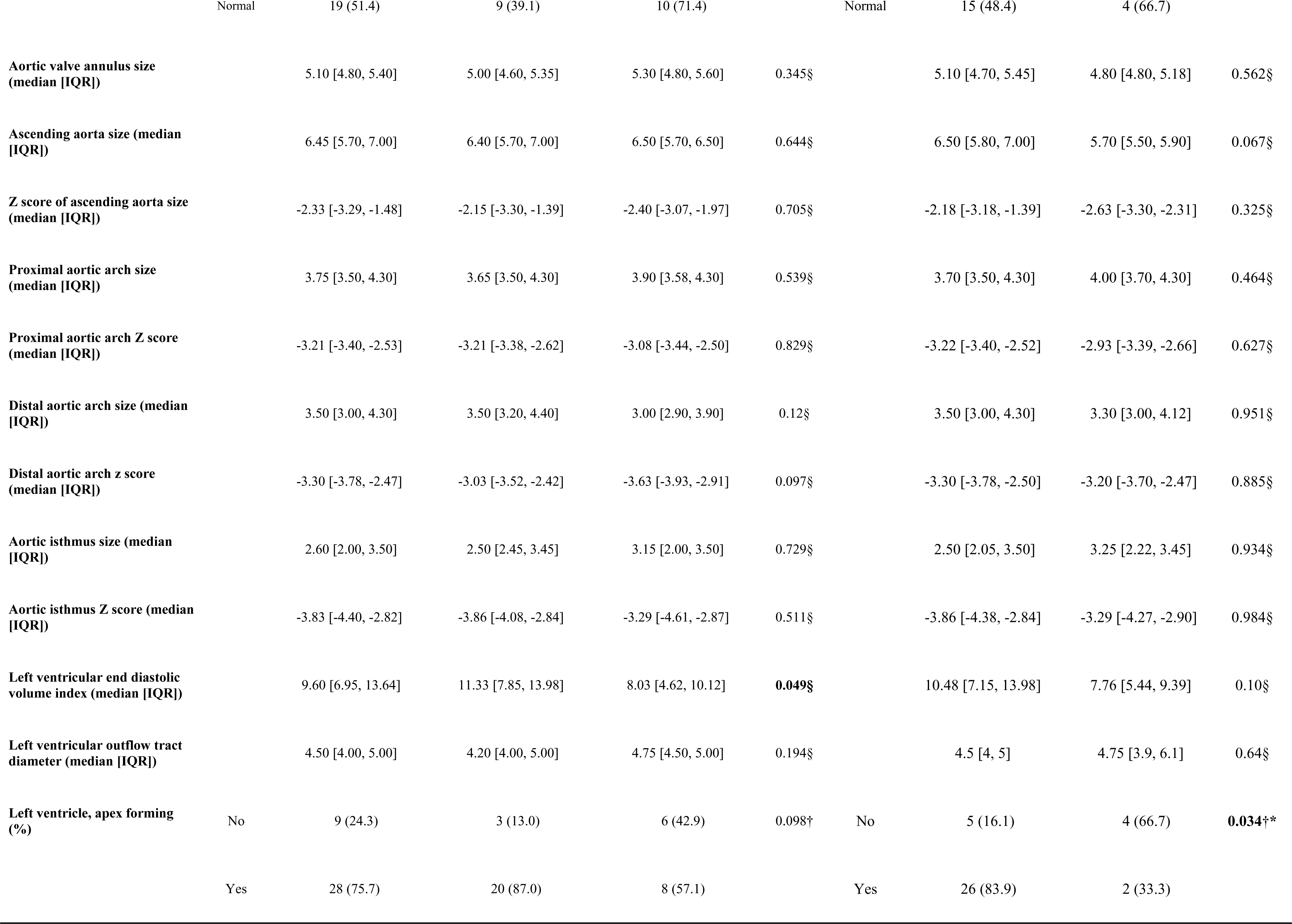
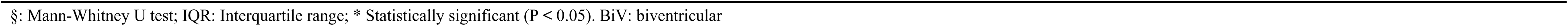
Summary of CMR and echo findings.

| Variable | Level | Overall | Primary Bi-V repair versus Hybrid procedure |  |  | Successful primary/staged Bi-V repair versus failed Bi-V repair |  |  |  |
| --- | --- | --- | --- | --- | --- | --- | --- | --- | --- |
|  |  |  | Successful Bi-V Repair<br>as Index Operation | Hybrid Initial Operation | P-value | Levels | Successful Primary Bi-V Repair as + Staged<br>Bi-V repair | Failure to achieve<br>Bi-V repair | P-value |
|  |  |  | (n = 23) | (n = 14) |  |  | (n = 31) | (n = 6) |  |
| CMR |  |  |  |  |  |  |  |  |  |
| Left ventricular mass index<br>(median [IQR]) |  | 22.00 [16.55, 24.60] | 23.50 [21.00, 25.80] | 17.00 [14.00, 22.00] | **0.049** *§ |  | 22.80 [16.92, 24.90] | 17.00 [13.70, 21.00] | 0.178§ |
| Left ventricular end diastolic<br>volume index (median [IQR]) |  | 28.00 [24.00, 30.00] | 29.00 [27.50, 31.00] | 20.00 [17.40, 25.75] | **<0.001** *§ |  | 28.00 [25.50, 31.00] | 17.40 [17.25, 18.60] | **0.002** *§ |
| Left ventricular end systolic<br>volume index (median [IQR]) |  | 12.00 [9.00, 14.00] | 13.00 [11.50, 14.00] | 8.30 [6.25, 10.75] | **0.001** *§ |  | 12.00 [10.50, 14.00] | 6.00 [5.70, 9.00] | **0.001** *§ |
| Left ventricular stroke volume<br>index (median [IQR]) |  | 16.00 [13.00, 18.00] | 17.00 [14.20, 18.00] | 11.70 [10.25, 13.45] | **<0.001** *§ |  | 16.00 [13.50, 18.00] | 11.40 [10.25, 13.15] | **0.016** *§ |
| Left ventricular ejection<br>fraction (median [IQR]) |  | 58.00 [50.00, 63.00] | 58.00 [51.50, 59.50] | 56.50 [50.50, 63.00] | 0.838§ |  | 57.00 [50.00, 59.50] | 63.00 [57.75, 66.75] | 0.122§ |
| Right ventricular end diastolic<br>volume index (median [IQR]) |  | 83.00 [67.60, 117.00] | 80.00 [60.50, 108.00] | 89.65 [81.50, 122.25] | 0.091§ |  | 83.00 [64.00, 113.00] | 82.50 [76.50, 114.00] | 0.636§ |
| Right ventricular ejection<br>fraction (median [IQR]) |  | 47.00 [42.00, 51.00] | 47.00 [41.50, 50.00] | 47.50 [43.00, 53.20] | 0.266§ |  | 46.50 [40.70, 50.00] | 56.30 [48.65, 59.75] | **0.021** *§ |
| Ascending aorta flow (median<br>[IQR]) |  | 1.90 [1.48, 2.23] | 2.10 [1.75, 2.36] | 1.27 [0.89, 1.81] | **<0.001** *§ |  | 1.99 [1.53, 2.28] | 0.97 [0.76, 1.69] | **0.012** *§ |
| Superior vena cava flow<br>(median [IQR]) |  | 1.36 [1.11, 1.60] | 1.40 [1.17, 1.60] | 1.33 [1.07, 1.88] | 0.695§ |  | 1.36 [1.11, 1.57] | 1.70 [1.24, 2.18] | 0.322§ |
| Ratio of ascending aorta to<br>superior vena cava flow<br>(median [IQR]) |  | 1.41 [1.00, 1.75] | 1.49 [1.27, 1.77] | 0.98 [0.60, 1.45] | **0.027** *§ |  | 1.44 [1.17, 1.77] | 0.85 [0.41, 1.29] | **0.034** *§ |
| Descending aorta flow (median [IQR]) |  | 1.05 [0.81, 1.40] | 1.10 [0.89, 1.38] | 1.00 [0.64, 1.42] | 0.38§ |  | 1.05 [0.80, 1.38] | 1.04 [1.00, 1.41] | 0.934§ |
| Left pulmonary artery flow (median [IQR]) |  | 1.50 [1.27, 2.62] | 1.50 [1.30, 2.71] | 1.62 [1.10, 2.32] | 0.594§ |  | 1.63 [1.30, 2.71] | 1.24 [1.10, 1.89] | 0.149§ |
| Right pulmonary artery flow (median [IQR]) |  | 2.29 [1.54, 3.41] | 2.15 [1.47, 3.45] | 2.38 [1.82, 3.38] | 0.719§ |  | 2.29 [1.61, 3.59] | 2.16 [1.61, 2.77] | 0.695§ |
| Ratio of pulmonary to systemic flow (median [IQR]) |  | 1.63 [1.05, 2.46] | 1.46 [1.04, 2.27] | 1.92 [1.18, 3.26] | 0.332§ |  | 1.95 [1.04, 2.88] | 1.46 [1.18, 1.76] | 0.458§ |
| Echocardiography |  |  |  |  |  |  |  |  |  |
| Left ventricular end diastolic dimension on short axis view (median [IQR]) |  | 13.20 [11.80, 14.20] | 13.90 [12.80, 14.25] | 12.30 [10.15, 13.17] | **0.015§** |  | 13.70 [12.30, 14.20] | 11.15 [9.88, 12.35] | **0.039§\*** |
| Z score of the left ventricular end-diastolic dimension (median [IQR]) |  | -3.74 [-4.71, -3.25] | -3.47 [-4.27, -3.04] | -4.41 [-5.52, -3.75] | **0.011§** |  | -3.66 [-4.53, -3.16] | -5.02 [-5.82, -4.22] | **0.032§\*** |
| Shortening fraction (median [IQR]) |  | 31.58 [28.20, 35.88] | 33.10 [29.47, 36.53] | 29.53 [24.95, 33.67] | 0.074§ |  | 32.39 [28.64, 35.92] | 31.30 [26.12, 33.10] | 0.257§ |
| Left ventricular length to right ventricular length ratio (median [IQR]) |  | 0.84 [0.81, 0.90] | 0.86 [0.83, 0.94] | 0.83 [0.75, 0.86] | **0.023§** |  | 0.85 [0.82, 0.92] | 0.78 [0.74, 0.85] | 0.143§ |
| Mitral valve 4 chamber view size (median [IQR]) |  | 6.90 [5.90, 7.30] | 7.00 [6.30, 7.50] | 5.75 [5.53, 6.75] | **0.006§** |  | 6.90 [6.15, 7.45] | 5.65 [5.35, 6.10] | **0.013§\*** |
| Z score of mitral valve size (median [IQR]) |  | -3.60 [-4.23, -3.08] | -3.26 [-3.75, -2.88] | -4.28 [-4.80, -3.84] | **0.001§** |  | -3.43 [-4.02, -2.92] | -4.51 [-5.10, -4.14] | **0.007§\*** |
| Mitral valve morphology (%) | Abnormal | 16 (43.2) | 8 (34.8) | 8 (57.1) | 0.322† | Abnormal | 13 (41.9) | 3 (50.0) | 1† |
|  | Normal | 21 (56.8) | 15 (65.2) | 6 (42.9) |  | Normal | 18 (58.1) | 3 (50.0) |  |
| Aortic valve morphology (%) | Bicuspid | 18 (48.6) | 14 (60.9) | 4 (28.6) | 0.117† | Bicuspid | 16 (51.6) | 2 (33.3) | 0.709† |
|  | Normal | 19 (51.4) | 9 (39.1) | 10 (71.4) |  | Normal | 15 (48.4) | 4 (66.7) |  |
| **Aortic valve annulus size (median [IQR])** |  | 5.10 [4.80, 5.40] | 5.00 [4.60, 5.35] | 5.30 [4.80, 5.60] | 0.345§ |  | 5.10 [4.70, 5.45] | 4.80 [4.80, 5.18] | 0.562§ |
| **Ascending aorta size (median [IQR])** |  | 6.45 [5.70, 7.00] | 6.40 [5.70, 7.00] | 6.50 [5.70, 6.50] | 0.644§ |  | 6.50 [5.80, 7.00] | 5.70 [5.50, 5.90] | 0.067§ |
| **Z score of ascending aorta size (median [IQR])** |  | -2.33 [-3.29, -1.48] | -2.15 [-3.30, -1.39] | -2.40 [-3.07, -1.97] | 0.705§ |  | -2.18 [-3.18, -1.39] | -2.63 [-3.30, -2.31] | 0.325§ |
| **Proximal aortic arch size (median [IQR])** |  | 3.75 [3.50, 4.30] | 3.65 [3.50, 4.30] | 3.90 [3.58, 4.30] | 0.539§ |  | 3.70 [3.50, 4.30] | 4.00 [3.70, 4.30] | 0.464§ |
| **Proximal aortic arch Z score (median [IQR])** |  | -3.21 [-3.40, -2.53] | -3.21 [-3.38, -2.62] | -3.08 [-3.44, -2.50] | 0.829§ |  | -3.22 [-3.40, -2.52] | -2.93 [-3.39, -2.66] | 0.627§ |
| **Distal aortic arch size (median [IQR])** |  | 3.50 [3.00, 4.30] | 3.50 [3.20, 4.40] | 3.00 [2.90, 3.90] | 0.12§ |  | 3.50 [3.00, 4.30] | 3.30 [3.00, 4.12] | 0.951§ |
| **Distal aortic arch z score (median [IQR])** |  | -3.30 [-3.78, -2.47] | -3.03 [-3.52, -2.42] | -3.63 [-3.93, -2.91] | 0.097§ |  | -3.30 [-3.78, -2.50] | -3.20 [-3.70, -2.47] | 0.885§ |
| **Aortic isthmus size (median [IQR])** |  | 2.60 [2.00, 3.50] | 2.50 [2.45, 3.45] | 3.15 [2.00, 3.50] | 0.729§ |  | 2.50 [2.05, 3.50] | 3.25 [2.22, 3.45] | 0.934§ |
| **Aortic isthmus Z score (median [IQR])** |  | -3.83 [-4.40, -2.82] | -3.86 [-4.08, -2.84] | -3.29 [-4.61, -2.87] | 0.511§ |  | -3.86 [-4.38, -2.84] | -3.29 [-4.27, -2.90] | 0.984§ |
| **Left ventricular end diastolic volume index (median [IQR])** |  | 9.60 [6.95, 13.64] | 11.33 [7.85, 13.98] | 8.03 [4.62, 10.12] | **0.049§** |  | 10.48 [7.15, 13.98] | 7.76 [5.44, 9.39] | 0.10§ |
| **Left ventricular outflow tract diameter (median [IQR])** |  | 4.50 [4.00, 5.00] | 4.20 [4.00, 5.00] | 4.75 [4.50, 5.00] | 0.194§ |  | 4.5 [4, 5] | 4.75 [3.9, 6.1] | 0.64§ |
| **Left ventricle, apex forming (%)** | No | 9 (24.3) | 3 (13.0) | 6 (42.9) | 0.098† | No | 5 (16.1) | 4 (66.7) | **0.034†\*** |
|  | Yes | 28 (75.7) | 20 (87.0) | 8 (57.1) |  | Yes | 26 (83.9) | 2 (33.3) |  |

§: Mann-Whitney U test; IQR: Interquartile range; \* Statistically significant ( $P < 0.05$ ). BiV: biventricular

#### 3.2.2 Echocardiographic Parameters

When compared to the hybrid/staging procedure group, patients who underwent a successful primary Bi-V repair, demonstrated larger absolute LVEDD (13.9 mm vs. 12.3 mm, p =0.015), larger LVEDD z-score (-3.47 vs. -4.41, p =0.011), larger LV volume (LVEDVi 11.33 mL/m^2^ vs 8.03 mL/m^2^, p = 0.049), larger MV dimension and z-score (7 mm vs. 5.75, p = 0.006; -3.26 vs. - 4.28, p = 0.001), and a larger LV/right ventricle (RV) length ratio (0.86 vs. 0.83, p = 0.023) ***(Table 2)***.

### 3.3 Imaging features associated with successful primary and successful staged Bi-V repair compared to failure to achieve Bi-V repair group

#### 3.3.1 CMR Parameters

Patients who underwent successful primary/ staged Bi-V repair exhibited higher values for LVEDVi (28 mL/ m^2^ vs. 17.4.00 mL/m2; p <0.002), LVSVi (16 ml/ m^2^ vs. 11.40 ml/ m^2^; p=0.016), higher blood flow volume through the ascending aorta (Q_AO_:1.99 L/min/ m^2^ vs. 0.97 L/min/ m^2^, p <0.012), and ascending aorta / superior vena cava (Q_SVC_) flow ratio (1.44 vs. 0.85, p =0.034) compared to those who had failure to achieve Bi-V repair (***Table 2***).

#### 3.3.2 Echocardiographic Parameters

When compared to those who had failure to achieve Bi-V repair, patients who underwent successful primary or staged Bi-V repair, demonstrated larger absolute LVEDD (13.7 mm vs. 11.15 mm, p =0.039), larger LVEDD z-score (-3.66 vs. -5.02, p =0.032), larger MV dimension and z-score (6.9 mm vs. 5.65, p = 0.013; -3.43 vs. -4.51, p = 0.007) ***(Table 2)***.There was no difference between the groups with regard to echo-derived LV volumes.

### 3.3 Comparison between echocardiography and CMR-derived left ventricular volumes

Echocardiogram and CMR-derived LVEDVi measurements were significantly correlated (r = 0.56, p <0.001) ***(Figure 4A)*** but showed poor agreement in Bland–Altman analysis with CMR-derived LVEDVi values being significantly larger than echo-derived LVEDVi (mean difference: 13.8±11.5; 95% CI: 36.1, -8.5) ***(Figure 4B)***.

**Figure 4.**
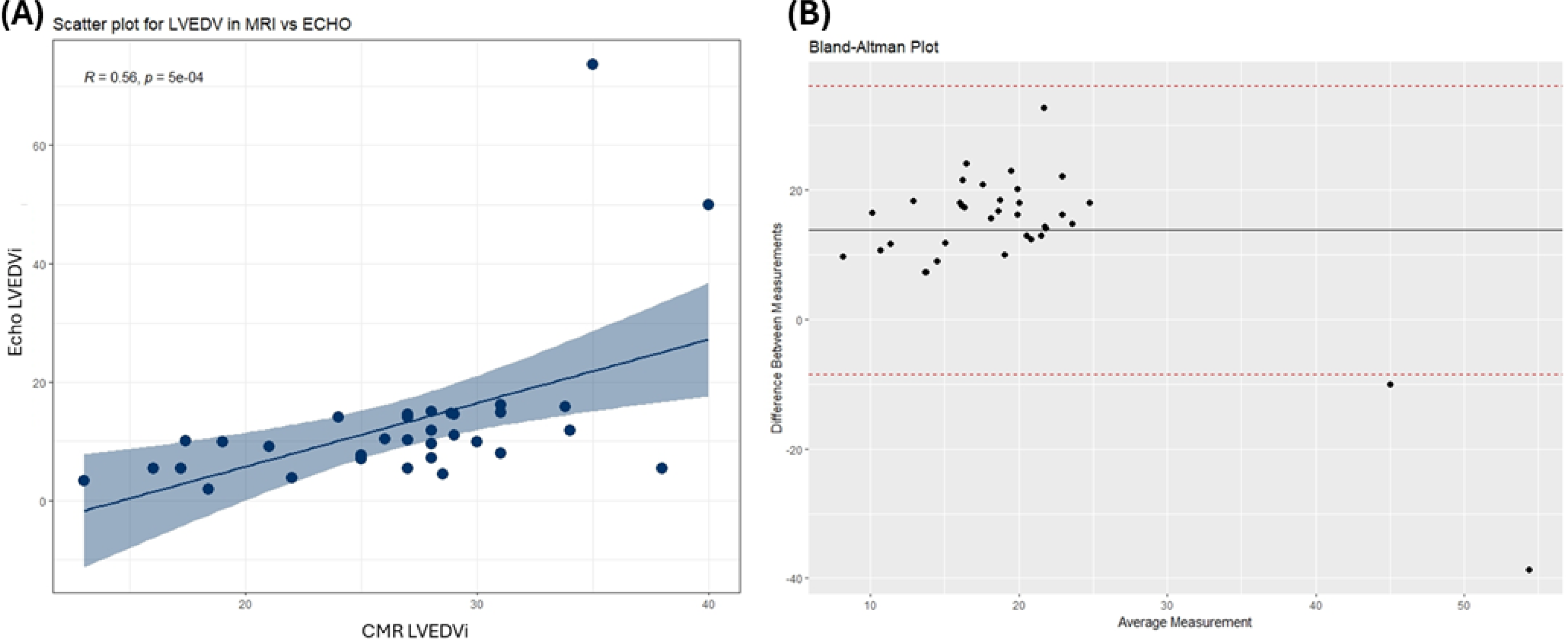
(A) Correlation between LVEDVi in MRI vs. Echo, (B) Bland-Altman plot between LVEDVi in MRI vs. Echo.

### 3.4 Diagnostic accuracy of CMR and Echo thresholds for primary successful Bi-V repair

Receiver operating curve (ROC) analysis was conducted to determine the CMR parameters and their corresponding threshold values that provided the most accurate prediction of successful primary Bi-V repair ***(Table 3, Figure 5)***. LVEDVi cutoff on CMR of 27 mL/m², had 87% sensitivity and 79% specificity with an AUC of 87.6% and Q_Ao_ threshold of 1.9 L/min/m^2^ had 65.2% sensitivity and 92.9% specificity (AUC: 86.0%). When ROC analysis was performed for echo parameters and threshold values that best predicted successful primary Bi-V repair, CMR parameters were found to be better predictors, with the best echo predictor being the z-score of MV size at -4.06 which had a sensitivity of 91.3%, specificity of 64.3%, and an AUC of 82.6%. Although echocardiogram and CMR-derived LVEDVi measurements were significantly correlated, echo-derived LVEDVi threshold of 21.2 mL/m^2^ had a lower AUC than CMR measured LVEDVi (74.2% vs 87.60%).

**Figure 5.**
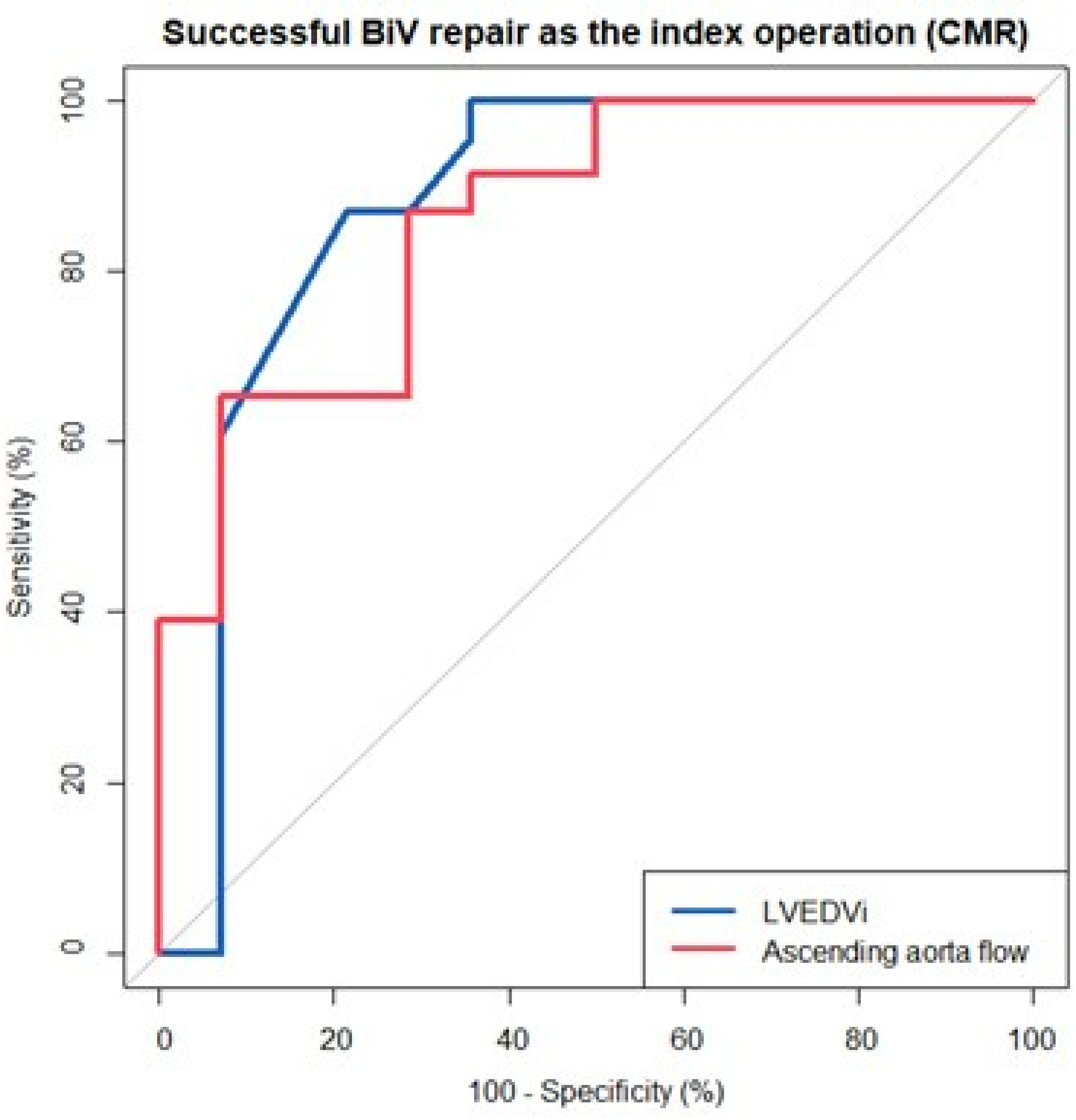
ROC curve for CMR in predicting successful Bi-V repair. LVEDVi cut off 27 m1/m2, 87% sensitivity and 79% specificity (AUC 87.6). QAO cut off of 1.99 L/min/m2, 65.2% sensitivity and 92.9% specificity (AUC 86.0%)

**Table 3.**
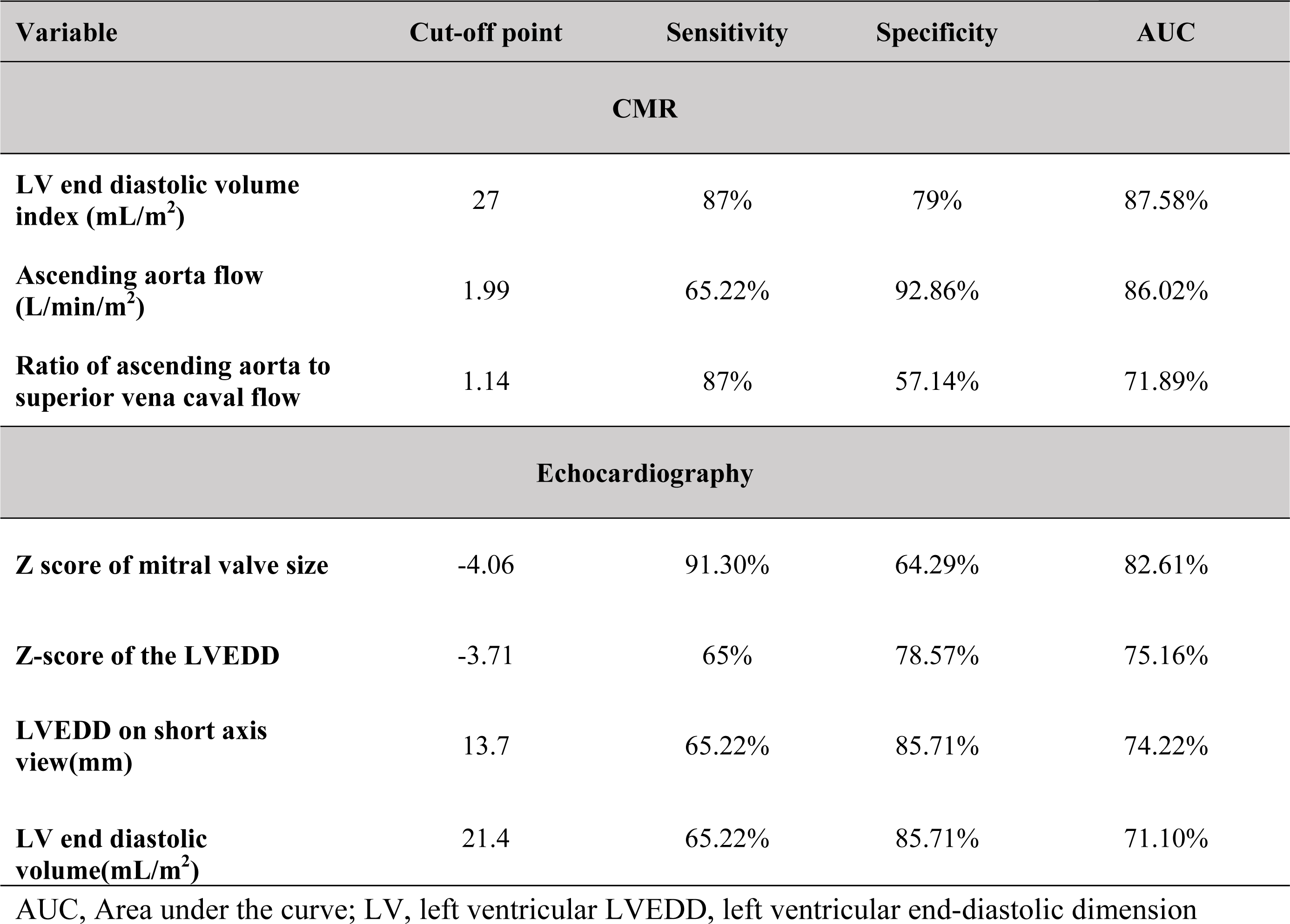
MRI and Echo thresholds for successful primary bi-ventricular repair.

### 3.5 Patient Outcomes

Twenty-three (62%) of the 37 patients underwent successful primary Bi-V repair and at a median follow-up of 9.44 (3.67-14.67) years, all patients remain alive. At the last follow up all patients had NYHA functional class 1/ Ross class 1, and 3 patients had moderate pulmonary hypertension. Among 8/37 (22%) who underwent successful staged Bi-V repair, median follow-up 4.3 (3.14-11.92) years, one patient had residual moderate MV stenosis and one had pulmonary hypertension. The remaining 6 (16%) patients with a median follow up 2.8(0.7-5.62) years failed to achieve a Bi-V repair (SV conversion, death/transplant) and four of these patients died, while one patient underwent heart transplantation. ***(Figure 1)***. Kaplan Meier curves for survival and freedom from reintervention showed no significant difference between the successful primary Bi-V repair and successful staged Bi-V repair (***Figure 6A and 6B)***.

**Figure 6.**
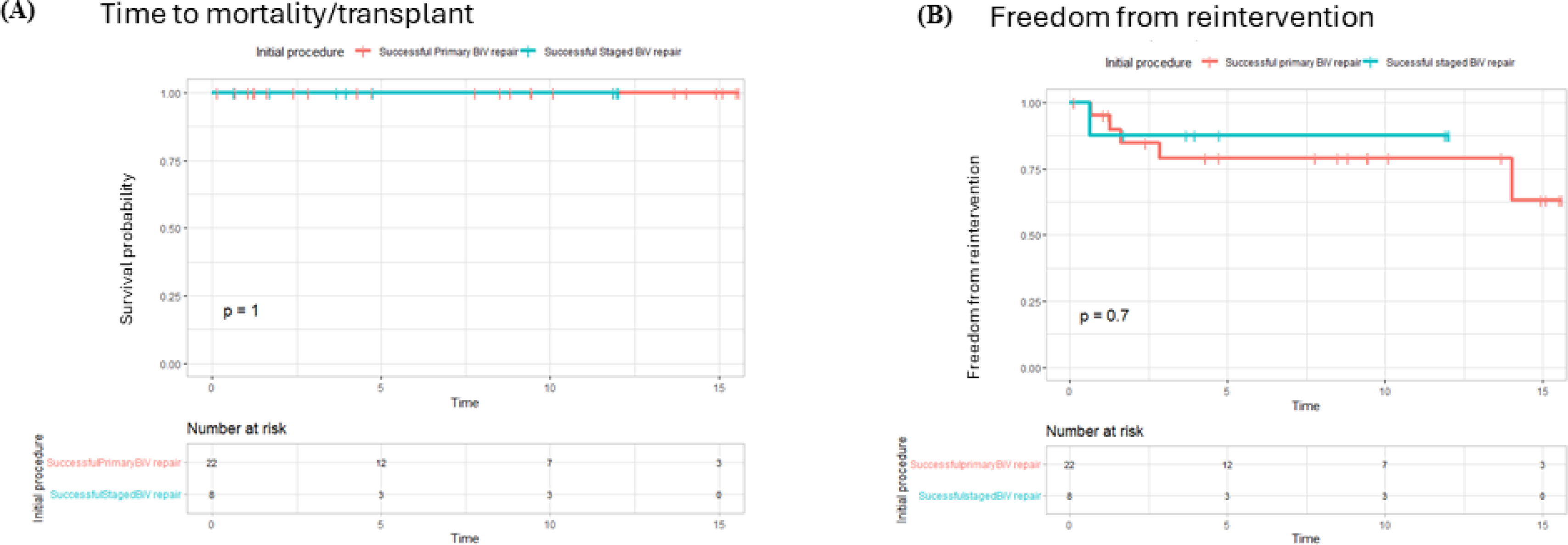
(A) Kaplan Meier Survival Curve of patients who underwent successful primary Bi-V repair and successful staged Bi-V repair. (B) Kaplan Meier curve of freedom from re-intervention of patients who underwent successful primary Bi-V repair and successful staged Bi-V repair.

#### 3.5.1 Comparison of imaging features between ‘successful staged Bi-V repair’ and ‘failure to achieve Bi-V repair group’

Within the hybrid/ staging procedure group (n=14), there were no significant differences in CMR-derived variables, including LVEDVi and Q_Ao_ between patients who had staged Bi-V repair or failure to achieve Bi-V repair (Supplementary Table 1). However, all patients with a failed Bi-V repair had initial LVEDVi <20 mL/m^2^. Follow-up echo data was available for all patients prior to definitive procedure (Bi-V/SV conversion), and CMR data was available for 10 / 14 prior to definitive procedure. Both groups demonstrated similar growth of LV size, mitral and aortic z-scores and, although interval increase in CMR-measured LVEDVi nearly missed statistical significance (p-value=0.06), there was no significant difference between both groups in terms of interval change of echocardiogram-derived LVEDD and LVEDD z-score, and MV and AV diameter or z-scores (Supplementary Table 2 and Supplementary Figure 1)

#### 3.5.2 Failure to achieve Bi-V circulation group

Of the 6 patients who failed to achieve Bi-V repair, 4 died, one underwent heart transplantation after staged Bi-V repair, and one patient remains alive following SV palliation. Specifically, the 4 patients who died had the smallest LV volumes of the entire cohort (13ml/ m^2^, 17.4 ml/ m^2^, 17.2 ml/m^2^ and 19 ml/m^2^). All 4 of these patients initially underwent a hybrid procedure, and demised despite undergoing SV palliation, after Stage 3 palliation (Fontan; n=1), comprehensive Stage 2 palliation (BCPS/Glenn n=2) and Norwood procedure (n=1).One patient underwent heart transplantation: this patient had initial LVEDVi of 17.4ml/m^2^, underwent staged Bi-V repair with arch repair, and closure of the ventricular septal defect at 6 months of age. This patient subsequently developed MV stenosis and LV outflow tract obstruction, needing MV repair and subaortic membrane resection. As this patient had persistent supra-systemic pulmonary hypertension, cardiac transplantation was performed at 1.6 years of age with resolution of pulmonary hypertension. The one surviving patient in this cohort with successfully competed SV palliation had an initial LVEDVi of 29 ml/m^2^ is doing well (Supplementary Table 3).

#### 3.5.3 Surgical interventions in Bi-V repair

23 patients achieved primary Bi-V and 8 patients achieved staged Bi-V repair. Of the 23 patients who underwent primary Bi-V repair, 15 patients underwent coarctation (CoA) repair, 1 patient underwent CoA repair with arch augmentation, 3 patients underwent CoA repair with arch augmentation and restriction of the atrial septal defect and 3 patients had CoA repair, arch augmentation and concomitant closure of the VSD with restriction/closure of the atrial septal defect. (Supplementary Table 4 and 5)

In the cohort of 31 patients who achieved a primary or staged Bi-V repair, 7 (22%) underwent reinterventions for significant residual lesions such as MV stenosis (5/7), multilevel LV outflow obstruction (5/7), AV stenosis (1/7) and re-coarctation (1/7) respectively (Supplementary Table 6). Four of the 7 patients had concomitant interventions for MV pathology and subaortic stenosis. A total of 21 reintervention procedures during the study period included 10 procedures for subaortic stenosis in 5 patients, 7 procedures for MV stenosis or supra-mitral ring in 5 patients, 3 procedures for AV stenosis in 3 patients, and 1 procedure for re-coarctation in 1 patient.

Subaortic stenosis requiring reintervention was due to progression of the unresected mild subaortic ridge in 2 patients and was newly developed in the narrow LV outflow tract in 3 patients. In 3 of these 5 patients, there was recurrence of subaortic stenosis after the surgical resection, requiring additional reintervention.

AV stenosis requiring surgical intervention was seen in 3 patients with bicuspid AV. In 2 of these 3 patients, aortic valve stenosis developed in the presence of recurrent subaortic stenosis. Mitral stenosis requiring reintervention was due to progression of subtle fibrous ridge on the atrial side of the MV annulus in 3 patients, progressive stenosis of the parachute MV in one, and unveiled stenosis of the MV with a papillary muscle arcade. Among these 5 patients, one patient developed severe pulmonary hypertension and received heart transplantation.

Note that *no* patients have required mitral valve replacement and there has been only one aortic valve replacement (Ross Procedure).

## 4. Discussion

Our study on patients with borderline LV hypoplasia in the absence of significant stenosis of aortic and mitral valves at the initial exam revealed several key findings. Firstly, we found higher CMR-derived LVEDVi, LVSVi, Q_Ao_, and a higher ratio of ascending aorta/SVC flow ratio to be significantly associated with successful primary and successful staged Bi-V repair. CMR derived LVEDVi threshold of ≥27 mL/ m^2^ predicted successful bi-V repair with 87% sensitivity and 79% specificity (AUC: 87.6%) to, while a CMR-derived Q_Ao_ threshold of 1.9L/min/ m^2^ predict successful Bi-V repair with a sensitivity of 65.2 % and specificity of 92.9 % (AUC: 86.00%). Further, 2D echocardiography consistently underestimated LV volume compared to CMR.

Current evidence to predict successful Bi-V repair in the specific cohort of patients with borderline LV hypoplasia without intrinsic valvar stenosis is limited, inconsistent, and predominantly based on echocardiography alone^14,15^. Mart and Eckhauser^15^ described an echocardiographically derived ‘2V-score’, a retrospectively derived echocardiographic score using a combination of measurements of MV, AV, LV and RV lengths, to predict successful Bi-V repair in this population. Similarly, Plymale^14^ et al described several echocardiographic features associated with successful Bi-V repair in a cohort of patients who underwent arch repair and had hypoplasia of the AV and/or MV. In their cohort of 73 patients, larger AV annulus and proximal transverse arch diameter were associated with favorable outcomes, while MV size was not associated with outcomes. Unlike them, others have reported^18,19^ that a morphologically abnormal and smaller MV ^20^ adversely impacted survival in these patients. Although it has been shown in our cohort and others ^21–24^ that echo underestimated LV volume, particularly when the LV is crescentic, few studies have evaluated the utility of CMR indices in patients with borderline LV hypoplasia without intrinsic stenosis of the AV and MV. Ours is the first study to evaluate CMR predictors of successful primary Bi-V repair in patients with borderline LV hypoplasia without significant stenosis of the mitral and aortic valves.

Based on our data, we recommend that in patients with borderline LV hypoplasia, decisions regarding functional LV adequacy in a Bi-V circulation should be based on CMR-derived LVEDVi which is the gold standard. Further based on ROC thresholds from our data, *primary Bi-V repair* can be safely considered in neonates with LVEDVi > 27 mL/m^2^ and Q_Ao_ > 1.9 L/min/m^2^. Kang et al^25^ reported that Q_Ao_ < 1.5 L/min/m^2^ portended worse survival and an increase in the hazard of failure of Bi-V circulation. Since ascending aorta flow depends on loading conditions, multi-level shunting, and left heart size, it could be viewed as an important measure that integrates both anatomy and physiology.

Further, in our cohort, almost all patients who had failed to achieve Bi-V repair had extremely small CMR-derived LV volumes, <20 mL/m^2^. Patients who first underwent a hybrid procedure and were then converted to SV palliation had a high mortality rate. This has been well described in other reports where pushing a patient with sub-optimal anatomy down the Bi-V pathway resulted in disproportionately higher morbidity and mortality.^8, 7^ As such in these patients LVEDVi <20 mL/m^2^, it may be prudent to pursue SV palliation at the outset to avoid risks of multiple interventions and the risk of developing hypoplasia of the branch pulmonary arteries from a prolonged period of banding.

Finally, patients with LVEDVi between 20 – 27 mL/ m^2^ who fall into a grey zone about the optimal primary management, should be considered candidates for the hybrid procedure as there is potential for LV catch-up growth. ^14,20,24^ In our cohort, all patients showed growth of LV structures during the interim period, although there was no statistically significant difference in LV growth between those who were able to achieve successful staged Bi-V repair compared to those who did not, this is likely due to the very small sample size (n=14). This further emphasizes the importance of CMR for follow-up imaging in these patients as it is a more accurate modality to assess LV size and growth.

While most patients in our cohort were able to achieve either primary or staged Bi-V repair, 22.5% of patients needed re-interventions within a few years after the initial procedure. This is comparable to re-intervention rates reported by Plymale et al^14^ (20%), and Grosse-Wortmann^24^ et al (16%).^24^ Others have reported higher reintervention rates with similar follow-up duration, with Freund et al^20^, reporting a reintervention rate of 37% and Mart & Eckhauser^15^ reporting a reintervention rate of 44%. This could reflect institutional practice and individualized thresholds for intervention but the reasons for reintervention cited in the literature are LV outflow tract stenosis and MV stenosis. Although surgical reintervention is a consistent feature in patients with borderline LV achieving Bi-V repair, it is important to note that no patients have required MV replacement and only one has required AV replacement (Ross Procedure). In our cohort also, LV outflow tract stenosis was the most common indication for reintervention. The reintervention data emphasizes early recognition of the substrate of subaortic stenosis and timely intervention as subaortic stenosis appears to prone to recur and the turbulent jet flow through the stenotic LV outflow may damage the AV leaflets causing aortic stenosis especially when the AV is bicuspid. The second most common indication for re-intervention was MV stenosis or a supra-mitral ring. The presence of a surgically amenable pathology such as a supra-mitral membrane or ring was identified in 3/ 5 patients, with others having a small MV annulus with failure to appropriately grow. Only one patient with progressive MV stenosis and worsening pulmonary hypertension needed heart transplantation, However, this patient had a very small LV to begin with (LVEDVi 17.4 mL/ m^2^ at birth) and a small MV (6.9 mm; z score -3.8). The very small LV at birth suggests, that this patient perhaps should have had SV palliation at the outset.

### Limitations

Our study had the inherent limitations of a retrospective review and a relatively small sample size. The immediate post-natal echocardiograms were obtained during the transitional neonatal circulation and may explain the smaller echocardiographic LV volumes when compared with CMR. The relatively small number of patients undergoing a hybrid procedure did not allow appropriate statistical analysis for comparison between cohorts with a Bi-V and SV outcome.

## Conclusions

CMR plays the critical role in pre-operative evaluation, surveillance and clinical decision-making in patients with borderline LV hypoplasia. In patients with borderline LV hypoplasia without significant stenosis of the MV and AV, successful primary biventricular repair can be achieved when the CMR-derived LVEDVi is greater than 27 mL/m^2^ and the ascending aorta flow is > 1.99 L/min/m^2^. In patients where the LVEDVi falls in the range of 20-27 ml/m^2^, a hybrid approach should be considered as a first intervention, which allow to defer the ultimate decision of biventricular versus single ventricle repair.

## Data Availability

All data referred referred to in the manuscript is available for review.

## ABBREVIATIONS

AV: aortic valve
Bi-V: Biventricular
CMR: Cardiovascular magnetic resonance
HLHC: Hypoplastic left heart complex
HLHS: Hypoplastic left heart syndrome
LV: Left Ventricle
LVEDD: Left ventricular end-diastolic dimension
LVEDVi: Left ventricular end diastolic volume index
LVSVi: Left ventricular stroke volume index
MV: mitral valve
Q_Ao_: Ascending aortic flow
Q_SVC_: Superior vena caval flow
RV: right ventricle
SV: Single ventricle
CoA: Coarctation

## Supplemental Material

Supplementary Tables 1–6, Supplementary Figure S1

